# A tool to distinguish viral from bacterial pneumonia

**DOI:** 10.1101/2021.06.22.21259301

**Authors:** Alfredo Tagarro, Cinta Moraleda, Sara Domínguez-Rodríguez, Mario Rodríguez, María Dolores Martín, María Luisa Herreros, María Dolores Folgueira, Alfredo Pérez-Rivilla, Julia Jensen, Agustín López, Arantxa Berzosa, Francisco José Sanz de Santaeufemia, Ana Belén Jiménez, Talía Sainz, Marta Llorente, Elisa Garrote, Cristina Muñoz, Paula Sánchez, Mar Santos, Marta Illán, Ana Barrios, Mónica Pacheco, Raquel Ramos Corral, Carmen Arquero, María Bernardino, Luis Prieto, Lourdes Gutiérrez, Cristina Epalza, Pablo Rojo, Lidia Oviedo, Miquel Serna-Pascual, Beatriz Soto, Sara Guillén, David Molina, Elvira Martín, Carmen Vázquez, Natalia Gerig, Cristina Calvo, María Pilar Romero, Manuel Imaz, Alfonso Cañete, José-Tomás Ramos, Juan Carlos Galán, Enrique Otheo, on behalf of VALS-DANCE Working Group.

**Affiliations:** Pediatrics Department. Hospital Universitario Infanta Sofía. Pediatrics Research Group. Universidad Europea de Madrid. Madrid, Spain; Pediatric Infectious Diseases Unit. Department of Pediatrics, Hospital Universitario 12 de Octubre; Pediatric Research and Clinical Trials Unit (UPIC). Instituto de Investigación Sanitaria Hospital 12 de Octubre (IMAS12), Madrid, Spain. Fundación para la Investigación Biomédica del Hospital 12 de Octubre, Madrid, Spain. RITIP (Traslational Research Network in Pediatric Infectious Diseases); Microbiology Department. Hospital Universitario Ramón y Cajal. Instituto Ramón y Cajal para la Investigación Sanitaria. Madrid, Spain; Microbiology Department. Laboratorio BR Salud. Hospital Universitario Infanta Sofía, San Sebastián de los Reyes, Madrid, Spain; Microbiology Department. Hospital Universitario 12 de Octubre. Instituto de Investigación Sanitaria Hospital 12 de Octubre (IMAS12), Madrid, Spain; Pediatrics Department. Hospital Infanta Cristina. Parla, Madrid, Spain; Pediatrics Deparment. Hospital Universitario Puerta de Hierro. Majadahonda, Madrid, Spain; Pediatrics Department. Hospital Universitario de Getafe. Getafe, Madrid, Spain; Pediatrics Department. Hospital Universitario Niño Jesús. Madrid, Spain; Pediatrics Department. Hospital Universitario Fundación Jiménez Díaz. Madrid, Spain; Pediatrics, Infectious and Tropical Diseases, Hospital Universitario La Paz. Instituto Investigación Hospital La Paz (IDIPAZ), Madrid, Spain. RITIP (Traslational Research Network in Pediatric Infectious Diseases); Pediatrics Department. Hospital Universitario del Sureste. Arganda del Rey, Madrid, Spain; Pediatrics Department. Hospital Universitario Basurto. Bilbao, Vizcaya, Spain; Pediatrics Department, Hospital General de Villalba. Villalba, Madrid, Spain; Pediatric Infectious Diseases, Immunology and Rheumatology Unit, University Hospital Virgen del Rocío. Instituto de Biomedicina de Sevilla (IBIS, Sevilla, Spain; Pediatric Infectious Diseases Unit. Hospital Universitario Gregorio Marañón. Madrid, Spain; Pediatrics Department. Hospital Clínico San Carlos. Madrid, Spain; Pediatrics Department. Universidad Complutense de Madrid. Madrid, Spain; Microbiology Department. Hospital Universitario de Getafe. Getafe, Madrid, Spain; Pediatrics Department. Hospital Universitario Ramón y Cajal; Microbiology Department. Hospital Universitario La Paz. Madrid, Spain; Microbiology Department. Hospital Universitario Basurto. Bilbao, Vizcaya, Spain; Universidad de Alcalá de Henares. Madrid, Spain

**Keywords:** Community-acquired pneumonia, typical bacteria, atypical bacteria, viral pneumonia

## Abstract

**Background and Objectives:** Establishing the etiology of community-acquired pneumonia (CAP) in children at admission is challenging. As a result, most children receive antibiotics that do not need.

This study aims to build and validate a diagnostic tool combining clinical, analytical and radiographical features to sequentially differentiate viral from bacterial CAP, and among bacterial CAP, typical from atypical bacteria, to improve choice of treatment.

**Methods:** Consecutive hospitalized children between 1 month and 16 years of age with CAP were enrolled. An extensive microbiological workup was performed. A score was built with a training set of 70% patients, to first differentiate between viral and bacterial CAP and secondly, typical from atypical bacterial CAP. To select variables, a Ridge model was used. Optimal cut-off points were selected to maximize specificity setting a high sensitivity (80%). Weights of each variable were calculated with a multivariable logistic regression. The score was validated with the rest of the participants.

**Results:** In total, 262 (53%) children (median age, 2 years, 52.3% male) had an etiological diagnosis.

The step 1 discriminates viral from bacterial CAP. Bacterial CAPs were classified with a sensitivity=97%, a specificity=48%, and a ROC’s area under the curve (AUC)=0.81. If a CAP was classificated as bacterial, it was assessed with step 2. The step 2 differentiates typical vs. atypical bacterial CAP. Typical bacteria were classified with a sensitivity=100%, a specificity=64%, and AUC=0.90.

**Conclusion:** This two-steps tool can facilitate the physician’s decision to prescribe antibiotics without compromising patient safety.

**Article summary:** We validated a clinical tool to predict the aetiology of CAP in children safely. This tool differentiates CAP into viral, atypical bacteria and typical bacteria.

**“What’s Known on This Subject”:** Establishing the aetiology of community-acquired pneumonia (CAP) in children at admission is challenging. As a result, most admitted children with CAP receive antibiotics.

**“What This Study Adds”:** We validated a clinical tool to predict the aetiology of pneumonia in children safely, differentiating among viral, atypical bacteria and typical bacteria.

## INTRODUCTION

Community-acquired pneumonia (CAP) is a significant cause of morbimortality worldwide. [1–3] Common aetiology are virus and bacteria. [4–6] However, when an individual patient is attended, aetiology is rarely achieved in real-time. Therefore, paediatricians have to decide empirically if a child needs antibiotics. As a result, most children receive antibiotics. [4,7]

We hypothesized that a two-steps score built from clinical and analytical features would differentiate most typical bacterial CAP accurately from viral and atypical bacterial CAP. The aim of this study was to build and validate a diagnostic tool to sequentially differentiate viral from bacterial CAP, and among bacterial CAP, typical from atypical bacteria.

## METHODS

### Study Design

This observational, multi-centre, prospective cohort study was conducted in two phases. The first pilot phase was performed at two hospitals in Madrid, Spain, from April 2012 to March 2015. The second phase was conducted in 15 hospitals in three regions of Spain (Madrid, País Vasco, and Andalucía), from December 2017 to May 2019.

Both phases were approved by the Ethics Boards of Hospital Universitario Ramón y Cajal (first phase, code 2011/0025) and Hospital 12 de Octubre (second phase, code 17/311) and the other participating hospitals. Informed consent was obtained from the guardians of all patients. Adapted information was given, and assent was obtained from patients from 12 to 16 years.

### Participants

Eligible participants were all children between 1 month and 16 years of age admitted to any of the participating hospitals, diagnosed as radiologically confirmed CAP, during the recruitment period. Enrolment was performed continuously until reaching a convenience sample of 150 participants in the first phase, and 300 participants in the second phase, plus a 10% of potential lost-to-follow-up. CAP was defined as fever and a compatible image in the chest X-ray (CXR) at admission. The interpretation of the CXR was performed following the standards of the “WHO Vaccine Trial Investigators Radiology Working Group”.[8] These standards establish 3 possible interpretations: “consolidation” (including consolidation and/or pleural effusion) and “other infiltrates”, or “normal”. Pleural effusion was confirmed with ultrasonography.

CAP was identified in the CXR by the attending paediatrician who admitted the participant and confirmed by radiologists at each centre. Exclusion criteria were the following: immunosuppressive conditions, suspected tuberculosis, chronic cardiac or pulmonary disease (except asthma), and hospital admission in the previous 30 days, and suspicion of lung aspiration or foreign body in the airway. Participants were followed-up until discharge.

### Microbiological Procedures

An extensive microbiological workup was performed. In short, we did blood cultures, *S. pneumoniae* antigen (BinaxNow™) and/or polymerase chain reaction (PCR) for *S. pneumoniae* in pleural fluid (PF) if thoracentesis was performed, PCR in blood for *S. pneumoniae*, PCR multiplex in nasopharyngeal aspirate (NPA) for pertussis and for 16 viruses: RSV, Human Metapneumovirus (hMPV), Parainfluenza virus (PIV) 1, 2, 3 and 4, Influenza (A and B), Human

Bocavirus (hBoV), Adenovirus (ADV), Enterovirus (EV), Rhinovirus (RhV), and Coronavirus

(CoV) 229E, OC43, NL63 and HKU12. The commercial systems xTAG® Respiratory Viral Panel Fast v1 (Luminex corp.) and CLART Pneumovir (Genomica SAU, Spain) were used. PCR for *M. pneumoniae* and *C. pneumoniae* was also performed using Mych Real Cycler-BIO-RAD CFX96, Progenie Molecular, Easy Mag (Biomérieux), and Mychle Real Cycler–BIO-RAD CFX96, Progenie Molecular. This system allows the qualitative detection by real-time PCR of the DNA of *M. pneumoniae* and *C. pneumoniae* in clinical samples. In all molecular tests, an internal extraction-amplification control was included to detect false negatives by PCR inhibition. Two paired samples for serology (at admission and 2-4 weeks afterward) of *M. pneumoniae* and *C. pneumoniae* was performed throughout enzyme immunoassay (ELISA) in 96-well plates, automated on Dynex platform and according to manufacturing companie’s protocols: Vircell, detection of IgG and IgM antibodies to *C. pneumonia* and detection of IgG antibodies to *M. pneumonia* and Palex Medical, detection of IgM antibodies to *M. pneumonia*).

### Definition of the etiological agent

The etiologic agents were defined according to the following criteria:

a. Likely typical bacterial infection: a bacterial pathogen (*S. pneumoniae, S. aureus, S. pyogenes, H. influenzae (Hib)*) detected in the blood through culture or PCR or, in PF, through culture, PCR or *S. pneumoniae* antigen detection.
b. Likely atypical bacterial infection: *M. pneumoniae* or *C. pneumoniae* detected by PCR in NPA or seroconversion or significative increase in IgG titles in the second sample, according to manufacturer instructions.
c. Likely viral infection: at least one putative pathogen respiratory virus (RSV, influenza A or B, PIV, hMPV) detected in NPA by PCR, and lack of a) or b). Other respiratory viruses (hRV, ADV, EV, COV, hBOV) were not included as likely viral infections due to poor specificity for CAP.[5,9,10]
d. In case of a positive putative virus detected in addition to a bacterium, CAP was classified as bacterial, since the final purpose of the study was to identify which patients would need antimicrobials.

### Databases

A manual selection based on clinical congruence of possible variables which predict the aetiology of pneumonia was made (n =18). They were radiographic image, gender, pneumococcal conjugate vaccine (PCV), influenza vaccine, fever days at admission, age at admission, vomiting, work of breathing (WoB), respiratory rate, maximum temperature, wheezing, oxygen saturation, leucocytosis >15 000 or leukopenia <4 000 cells/mm^3^, neutrophilia > 10 000 cells/mm^3^, sodium, haemoglobin, and C-reactive protein (CRP). Subjective variables or difficult to collect were excluded.

### Statistical Analysis

The categorical variables were presented as frequency distributions and the continuous variables were presented as median and interquartile ranges (IQR). To assess differences, we performed a chi-squared test for categorical variables and the Mann-Whitney *U* test for continuous variables.

The complete dataset was randomly split into a training set with 70% of the registers (n=184) and the remaining 30% for testing (n=78). This partition was balanced based on the aetiology (bacterial, viral).

The predictive relative variable importance for predicting bacterial and typical bacterial aetiology was assessed by a Ridge regression model. The variables with more than 10% of relative importance were selected to be included in the score.

For each of the steps of the score, the selected continuous variables were categorized using the optimal bootstrapped cut-off points selected by maximizing specificity while maintaining a sensitivity above 80% for detecting bacterial aetiology.

The score was built using two multivariable logistic models. In the first step, we extracted the odds ratio for variables associated with bacterial aetiology (ref: viral aetiology) in the training set. Afterwards, we selected only the patients with bacterial aetiology from the training set (n=87) and extracted the odds ratio for variables assessing the risk of typical bacterial compared to atypical bacterial aetiology. Those variables with few outcome events per level and/or large odds ratio with a wide confidence interval (infinite or +1000) were excluded to avoid sparse data bias. Finally, the total score was calculated for each subject to represent the prediction of the aetiology probability. The optimal cut-point of each step was selected by maximizing specificity while maintaining sensitivity above 80% for bacterial aetiology and typical bacterial aetiology, respectively. Both steps of the score were externally validated in the testing dataset. The performance of each score was assessed describing the sensitivity, specificity, accuracy, and AUC.

The missing values of both partitions (training/testing) were imputed using a non-parametric algorithm based on random forest. The normalized root mean squared error (NRMSE) and the proportion of falsely classified (PFS) was assessed for continuous and categorical variables, respectively. All the statistical analyses were performed using the R language.

### Web app

The score formula was implemented into a decision support tool app to make the aetiology classification comprehensive, easy and friendly to the physicians. The app provides probability of aetiology, and the user should interpret it as a guide for treatment. The app is available at https://saradominguez-rodriguez.shinyapps.io/ValsDance_app/ (username: user, password: 0000).

## RESULTS

A total of 495 children were enrolled, 151 in phase 1 and 344 in phase 2. Of them, 465 (94%) received antibiotics at admission, and 371 (74.9%) completed all the tests and the follow-up. At least a likely causative pathogen was identified retrospectively in 262 patients (52.9%). A total of 138 (52.7%) were attributed to viral aetiology and 124 (47.3%) to bacterial aetiology. Of them, 40 (15.3%) were attributed to typical bacterial aetiology, and 84 (32.1%) were attributed to atypical bacteria (Table 1).

The predictors included in the first step of the score, which aims to classify bacterial from viral aetiology are displayed in Table 2 and plotted by importance in Supplementary Figure 1.

According to the optimal cut-off point, age at admission was categorized as ≥3 years for both steps, haemoglobin was categorized as ≥11 g/dL in both score steps, and maximum temperature was categorized as ≥37.7ºC in the step 1.

The predictors included in the second step, which aims to classify typical from atypical bacterial aetiology are also displayed in Table 2 and plotted by importance in Supplementary Figure 2.

Step 1 (viral CAP vs. bacterial CAP). The weights for each level and variable of the score were calculated from the odds ratios in the multivariable model (Supplementary Figure 3). The step 1 discriminated bacterial CAP using the information of: CXR (consolidation, +5.5 points), age at admission (≥3y, +10.6), WoB (lack of WoB, +2.2), wheezing (no wheezing, +1), temperature (≥37.7ºC, +1.3), PCV (0 doses, +1.2), leucocytosis >15 000 cells/mm^3^ or leukopenia <4 000 cells/mm^3^ (+1.1), neutrophilia >10 000 cells/mm^3^ (+1.2), haemoglobin (≥11 g/dL, +2.3), CRP (>100 mg/dL, 2.2). The sum of the weights for each patient was calculated to know the score of each patient. The optimal cut-off point for the step 1 to classify a CAP with high sensitivity for bacterial aetiology was ≥11 points (sensitivity 93.1%, specificity 57.7%, AUC=0.80, Figure 1). In the external validation, bacteria were classified with a sensitivity 97.3%, specificity 48.8%, positive predictive value 63.2%, negative predictive value 95.2%, and AUC=0.81 (Supplementary figure 4).

**Figure 1.**
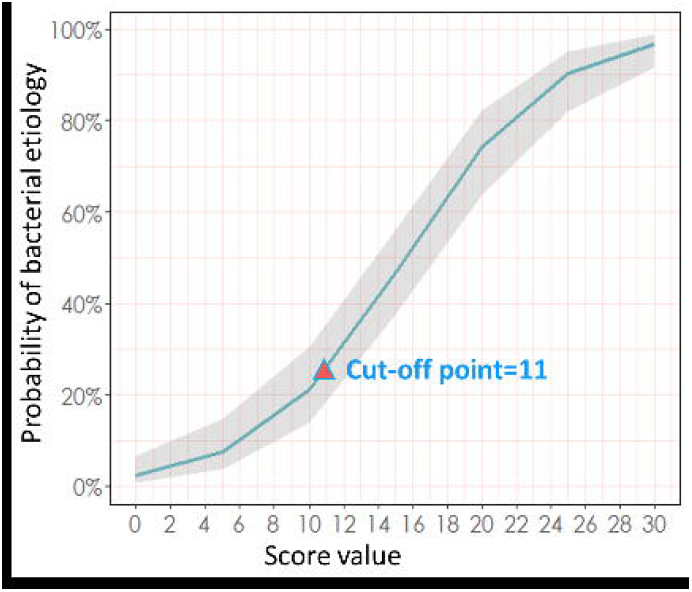
Probability of bacterial aetiology according to the results of the sum of values of Step 1. A significant risk of having a typical bacterial pneumonia was set on 11. Children with >11 points have >25% risk of bacterial pneumonia and paediatricians should consider prescription of antibiotics. Below 11 points, the risk of bacterial pneumonia is below 25%. Antibiotics directed against bacterial pneumonia may not be necessary. For an optimal choice of antibiotics, step 2 can be informative (see Figure 2).

### Step 2 (atypical bacteria vs. typical bacteria)

In step 2, participants who scored as bacterial in step 1 were included. According to the multivariable model the step 2 was built with: age at admission (<3 years, +6.8), cough (no, +3), wheezing (no wheezing, +5.0), WoB (yes, +5.8), haemoglobin (<11 g/dL, +5.4), leucocytosis >15 000 cells/mm^3^ or leukopenia <4 000 cells/mm^3^ (+2.4), and neutrophilia >10 000 cells/mm^3^ (+3.3). The sum of the weights for each patient was calculated to know the step 2 of each patient. The CRP (OR: 14.5 [3.1-86.9], p=0.001), influenza virus vaccine (OR: 2.3·10^8^ [3.2·10^−147^-Inf], p=0.994), and radiograph image interpretation (OR: 1.3·10^8^ [2.4·10^−54^-Inf], p=0.992) were excluded in the final model due to their wide confidence interval (Supplementary figure 4). The optimal cut-off points for the step 2 to classify a CAP as of typical bacterial aetiology was ≥11.7 points (sensitivity 93.3%, specificity 61.4%, AUC=0.89). Atypical bacteria aetiology was classified with <11.7 points (Figure 2). In the validation, typical bacteria were classified with a sensitivity 100%, specificity 64%, positive predictive value 37%, negative predictive value 100%, and AUC=0.90 (Supplementary figure 5). The distribution of the patients with identified aetiology across the two steps is displayed in Figure 3.

**Figure 2.**
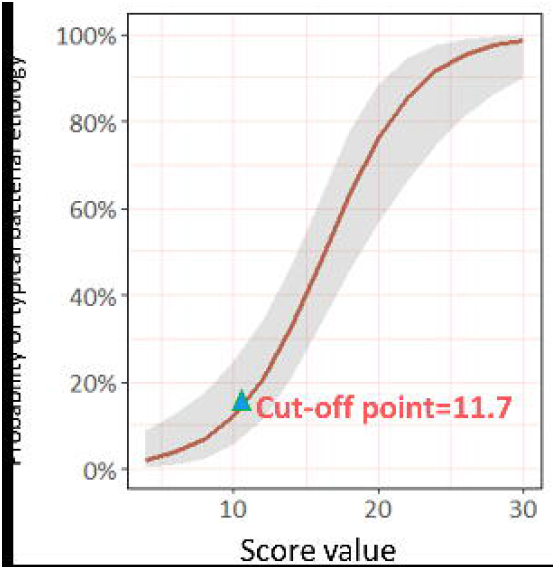
Probability of typical bacterial etiology according to the results of the sum of values of Step 2. A significant risk of having a typical bacterial pneumonia was set on 11.7. Children with at least 11.7 points have >18% risk of typical bacterial pneumonia and should receive antibiotics specifically directed against typical bacteria. Below 11.7 points, the risk of typical bacterial pneumonia is below 18%. Antibiotics directed against typical bacteria may not be necessary. Antibiotics directed against atypical bacteria might be considered.

**Figure 3.**
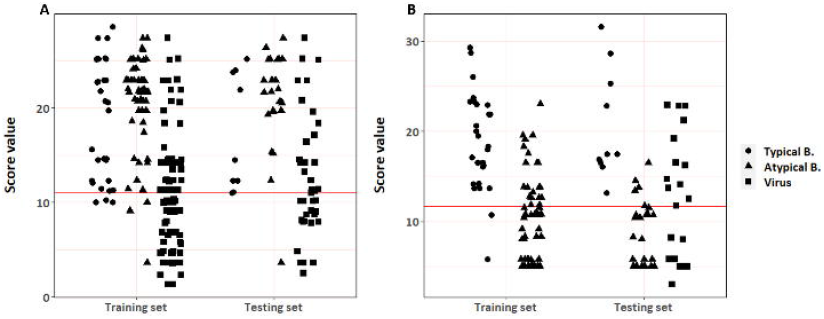
Distribution of the sample, according to likely etiologies and results of all patients in step 1 and step 2. A: Step 1. Only 3/29 (10.3%) typical bacteria scored as viral, and 2/50 (4 %) atypical bacteria scored as viral in the training set. No typical bacteria and only 1/34 (3%) atypical bacteria scored as viral in the testing set. This patient had PCR positive for *M. pneumoniae* and human metapneumovirus in the nasopharyngeal aspirate. No serial serologies were available. B: Step 2. Only 2/29 (7%) typical bacteria scored as atypical in the training set. No typical bacteria scored as atypical in the testing set. 4/34 (12%) atypical bacteria scored as typical in the testing set.

## DISCUSSION

In this study, we propose that most viral, typical bacterial and atypical bacterial CAPs can be differentiated at the time of admission with a score built from easily available clinical, radiographic, and analytical parameters. The use of this score can be facilitated by an online app. The online app provides probabilities of bacterial infection and, among them, typical or atypical bacterial infection.

Some features traditionally considered reliable markers of typical pneumonia, as consolidation and high CRP, were not included in step 2. The reason is that the number of events of “other infiltrates” or low CRP was too sparse to estimate the risk, so the confidence interval (CI95%) was too wide and the certainty was low. Some studies have suggested that PCT has good accuracy for differentiating RSV from *S. pneumoniae* CAP or viral from bacterial CAP [11–14]. PCT and albumin were included in the protocol but were not used in the model due to missing data.

Some of the items that predicted typical bacterial aetiology are not so obvious, like haemoglobin. Haemoglobin is not a classical marker of viral/bacterial infection, but inflammation is an important cause of anaemia which explains the association of anaemia with typical bacteria showed in the step 2.

In previous research, wheezing and CXR with “other infiltrates” have been suggested as predictors of viral CAP [15], but the distinction between viral and atypical bacteria, and typical from atypical bacteria is not so straightforward, because of significant overlapping. [16,17]

We suggest that patients who score below 11 in step 1 could be safely treated without antibiotics. Around half of antibiotics that were used for children with viral CAP had been saved with this tool (Supplementary figure 6) No typical bacteria and only 1/34 (3%) atypical bacteria scored as viral in the testing set of step 1. In addition, no typical bacteria scored as atypical in the testing set of step 2. Only 4/34 (11.8%) atypical bacteria scored as typical in the testing set. However, this is not considered of high relevance since the benefit of antibiotics for atypical bacteria is controversial. Some patients with high score in step 1 had only virus detected. We hypothesize that these patients may have undetected bacterial coinfections, or that they developed superinfections with bacteria. If these patients actually had a bacterial infection as expected, the accuracy of the score would be even better than reported.

The value and novelty of this tool are their high predictive values. Some scores tried to achieve the same aim as we did, but the microbiological gold standards were less accurate.[18],[19] With this tool, we can safely save many antibiotics routinely used for CAP in children, which may have an impact on the antimicrobial stewardship.

One of the main limitations of this study is the low specificity of the scores. We prioritized sensitivity over specificity in order to avoid misdiagnosis of bacteria in the first step and typical pneumonia in the second, because CAP caused by typical bacteria are potentially the most severe, have more complications, and are treatable. Therefore, a CAP with ≥25% probability of being caused by typical bacteria is classified by this tool as caused by typical bacteria to prevent important false negatives. We considered unacceptable the risk of not treating with antibiotics against typical bacterial a child with ≥25% probability of a serious typical bacterial infection. The well-known and inherent poor sensitivity of the current methods to identify bacterial infections limits the certainty of bacterial attribution. Therefore, we had to compare our scores to imperfect gold standards. In research where gold-standard is not clear, test accuracy indexes should not be taken as a hard fact. However, the microbiological approach we used is close to the best available gold standard in clinical practice.

Reproducibility of these results should be explored in different settings, especially in areas without routine immunizations for *S. pneumoniae* and *H. influenzae*, or where cut-off values for Hb or other parameters may be different. This analysis was performed before the COVID-19 pandemic. SARS-CoV-2 should be ruled out prior to using this tool.

## CONCLUSION

We provide a validated clinical tool to differentiate viral, typical, and atypical CAP safely. This tool can improve the appropriate use of antibiotics in paediatric CAP.

## Data Availability

On request after consideration

## Acknowledgments / Other members of VALS DANCE Working Group

Rut del Valle (Hospital Universitario Infanta Sofia), Julia Yebra (Hospital Universitario Infanta Sofia), Rosa Batista (Hospital Universitario Infanta Sofia), Teresa Raga (Hospital Universitario Infanta Sofia), Maria García-Baró (Hospital Universitario Infanta Sofia), Magdalena Hawkins (Hospital Universitario Infanta Sofia, Universidad Europea), Daniel Blázquez (Hospital Universitario 12 de Octubre), Manuel Gijón (Hospital Universitario 12 de Octubre), Lucía Figueroa (Fundación para la Investigación Biomédica del Hospital 12 de Octubre), Nazaret del Amo (BR Salud, Hospital

Universitario Infanta Sofía), Ana Méndez-Echeverría (Hospital Universitario La Paz), Mercedes Alonso-Sanz (Hospital Universitario Niño Jesús), Esther Casado Verrier (Hospital de Villalba), María José Cilleruelo (Hospital Universitario Puerta de Hierro), María Luz Golmayo (Hospital Universitario Puerta de Hierro), María Isabel Sánchez (Hospital Universitario Puerta de Hierro), Teresa del Rosal (Hospital Universitario La Paz), Alfonso Rodríguez-Albarrán (Hospital de Arganda).

We thank all the patients and families for their participation in this cohort, and the staff members who cared for them.

## Abbreviatures

ADV: Adenovirus
AUC: Area under the curve
CAP: Community-acquired pneumonia
CoV: Coronavirus
CRP: C-reactiva protein
CXR: Chest X-ray
EV: Enterovirus
hBoV: Human Bocavirus
hMPV: Human Metapneumovirus
NPA: Nasopharyngeal aspirate
PCV: pneumococcal conjugate vaccine
PIV: Parainfluenza virus
PCR: Polymerase Chain Reaction
PCT: Procalcitonin
ROC: Receiver Operator Curve
RhV: Rhinovirus
RSV: Respiratory Syncitial Virus
WoB: work of breathing

## FIGURE LEGENDS

**Supplementary figure 1.**
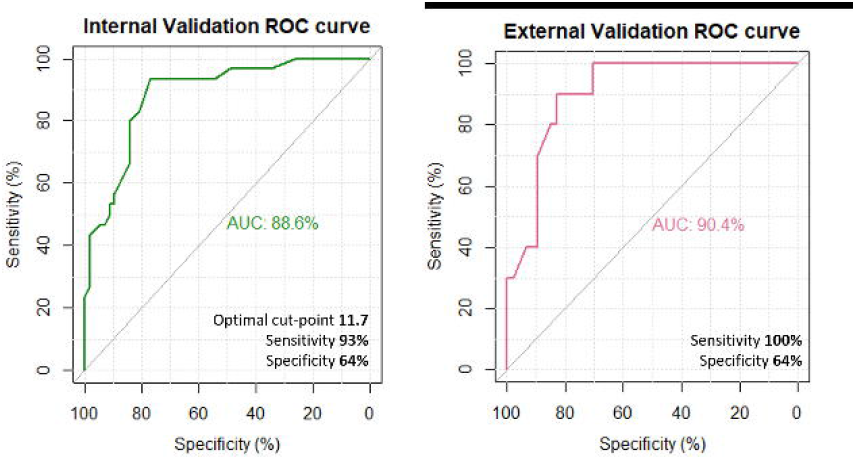
The ten most relevant variables to differentiate typical from atypical bacteria-associated community-acquired pneumonia.

**Supplementary figure 2.**
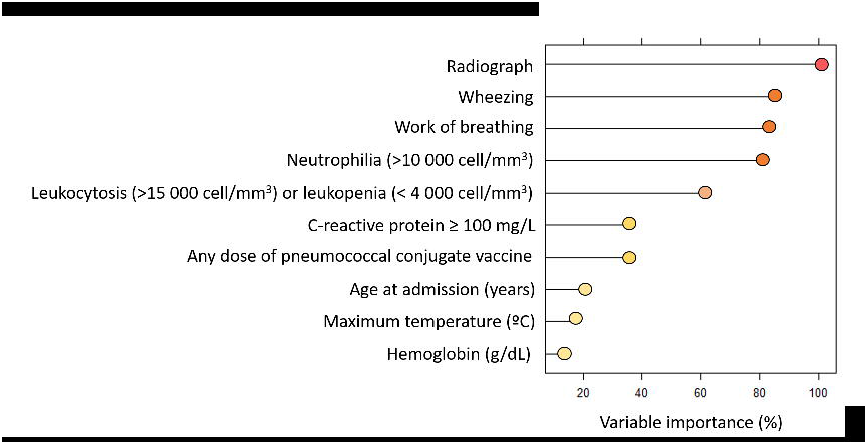
The ten most relevant variables to differentiate viral from bacterial community-acquired pneumonia.

**Supplementary figure 3.**
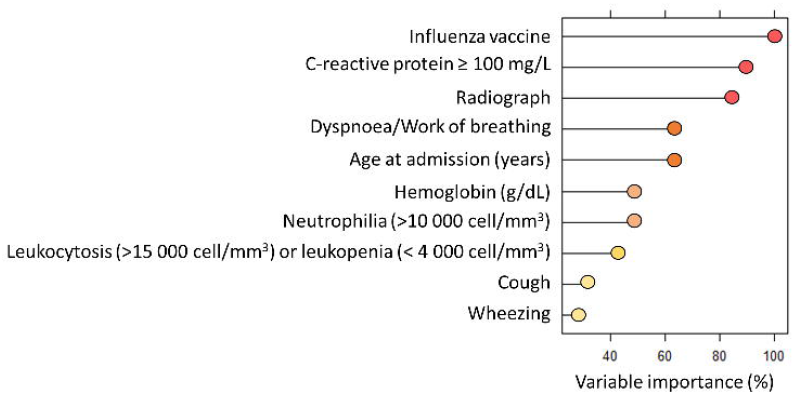
Multivariant model for step 1 to differentiate viral from bacterial community-acquired pneumonia. * p<0.050; ** p<0.01; ***p<0.001.

**Supplementary Figure 4.**
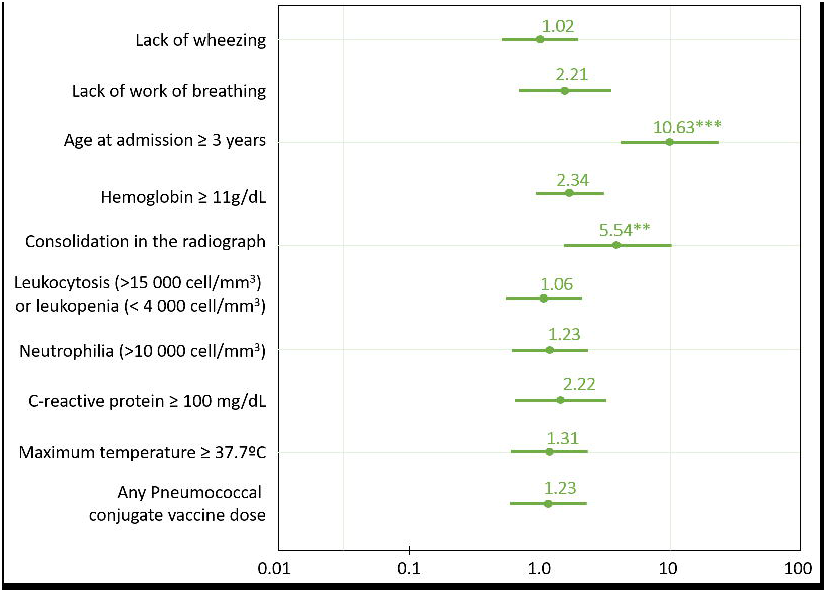
Multivariant model for step 2 to differentiate typical bacterial from atypical bacterial community-acquired pneumonia. C-reactive protein, influenza vaccine, and consolidation in the radiograph were excluded in the model because of their large odds ratio with a wide confidence interval (CI95%) to avoid sparse data bias (see results for the odds ratio and CI95% of C-reactive protein, influenza vaccine and consolidation in the radiograph). * p<0.050; ** p<0.01; ***p<0.001.

**Supplementary figure 5.**
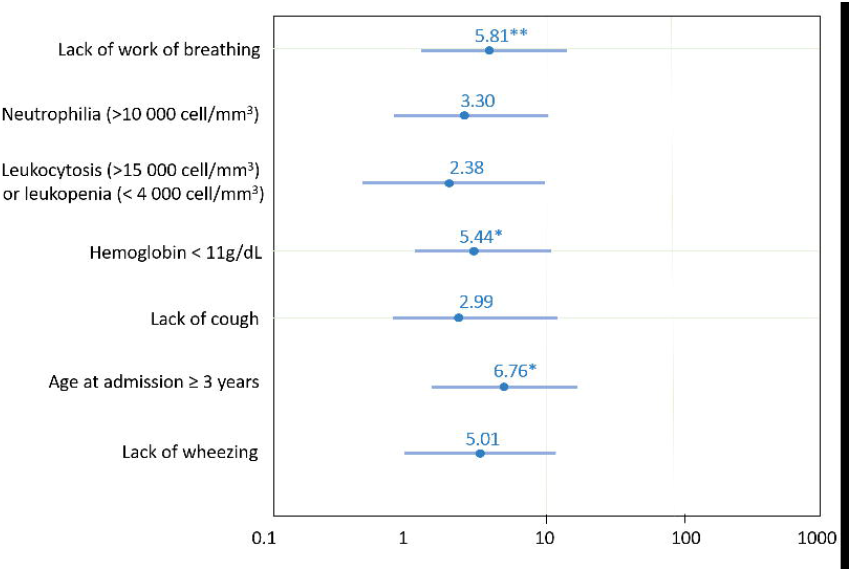
Validation of step 1. External validation refers to the testing set, after internal validation in the training set. AUC: Area under the curve.

**Supplementary figure 6.**
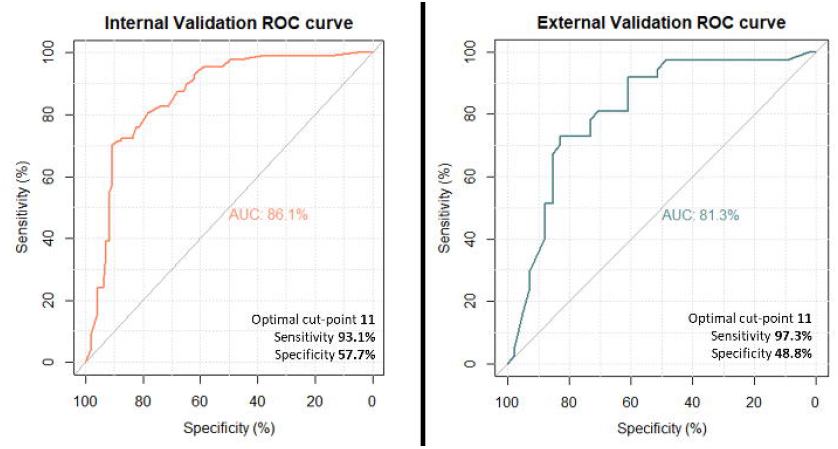
Validation of Step 2. External validation refers to the testing set, after internal validation in the training set. AUC: Area under the curve.

