## Supplementary material for "A tool to distinguish viral from bacterial pneumonia": Table 1

**Table 1. Features of patients according to training and testing groups.**

|  | **[ALL]** | **Test** | **Train** | **p.overall** |
| --- | --- | --- | --- | --- |
|  | ***N=262*** | ***N=78*** | ***N=184*** |  |
| Age at admission (years) -  Median (IQR) | 2.00 [1.00;5.00] | 2.00 [1.00;4.00] | 3.00 [1.00;5.00] | 0.338 |
| Gender |  |  |  | 0.463 |
| Male – n (%) | 137 (52.3%) | 44 (56.4%) | 93 (50.5%) |  |
| Female– n (%) | 125 (47.7%) | 34 (43.6%) | 91 (49.5%) |  |
| Influenza vaccine |  |  |  | 1.000 |
| Yes – n (%) | 13 (4.96%) | 4 (5.13%) | 9 (4.89%) |  |
| No – n (%) | 249 (95.0%) | 74 (94.9%) | 175 (95.1%) |  |
| Days of Fever - Median (IQR) | 4.00 [2.00;5.00] | 4.00 [2.00;6.00] | 4.00 [2.00;5.00] | 0.277 |
| Vomits |  |  |  | 0.196 |
| Yes – n (%) | 97 (37.0%) | 34 (43.6%) | 63 (34.2%) |  |
| No – n (%) | 165 (63.0%) | 44 (56.4%) | 121 (65.8%) |  |
| Maximum Temperature Median (IQR) | 39.0 [38.1;39.6] | 39.0 [38.4;39.5] | 39.0 [38.1;39.6] | 0.808 |
| Pneumococcal Conjugate Vaccine |  |  |  | 0.156 |
| Any doses – n (%) | 245 (94.2%) | 71 (91.0%) | 174 (95.6%) |  |
| Zero doses – n (%) | 15 (5.77%) | 7 (8.97%) | 8 (4.40%) |  |
| Respiratory frequency Median (IQR) | 38.0 [28.0;48.0] | 40.0 [30.2;50.0] | 37.0 [28.0;45.5] | 0.107 |
| Work of Breathing |  |  |  | 0.152 |
| No – n (%) | 82 (31.3%) | 19 (24.4%) | 63 (34.2%) |  |
| Yes – n (%) | 180 (68.7%) | 59 (75.6%) | 121 (65.8%) |  |
| Wheezing |  |  |  | 0.940 |
| Yes – n (%) | 100 (38.2%) | 29 (37.2%) | 71 (38.6%) |  |
| No – n (%) | 162 (61.8%) | 49 (62.8%) | 113 (61.4%) |  |
| Oxygen saturation (%) - Median (IQR) | 94.0 [91.0;96.0] | 94.0 [92.0;96.0] | 94.0 [91.0;96.0] | 0.986 |
| Radiograph |  |  |  | 0.504 |
| Consolidation – n (%) | 203 (77.5%) | 63 (80.8%) | 140 (76.1%) |  |
| Other infiltrates – n (%) | 59 (22.5%) | 15 (19.2%) | 44 (23.9%) |  |
| Hemoglobin - Median (IQR) | 11.9 [10.9;12.7] | 11.5 [10.9;12.4] | 12.1 [11.0;12.9] | 0.032 |
| Lymphocytes - Median (IQR) | 2400 [1500;3822] | 2425 [1564;4025] | 2400 [1400;3808] | 0.720 |
| Leukocytosis or Leukopenia |  |  |  | 1.000 |
| No – n (%) | 172 (65.6%) | 51 (65.4%) | 121 (65.8%) |  |
| Yes – n (%) | 90 (34.4%) | 27 (34.6%) | 63 (34.2%) |  |
| Neutrophilia |  |  |  | 1.000 |
| No – n (%) | 173 (66.0%) | 52 (66.7%) | 121 (65.8%) |  |
| Yes – n (%) | 89 (34.0%) | 26 (33.3%) | 63 (34.2%) |  |
| C-reactive protein ≥ 100 mg/L |  |  |  | 0.262 |
| No – n (%) | 163 (62.2%) | 44 (56.4%) | 119 (64.7%) |  |
| Yes – n (%) | 99 (37.8%) | 34 (43.6%) | 65 (35.3%) |  |
| Procalcitonin ≥ 1.5 ng/mL |  |  |  | 0.748 |
| No – n (%) | 155 (74.5%) | 47 (72.3%) | 108 (75.5%) |  |
| Yes – n (%) | 53 (25.5%) | 18 (27.7%) | 35 (24.5%) |  |
| Sodium - Median (IQR) | 137 [135;138] | 136 [134;138] | 137 [135;139] | 0.138 |
| Hypoalbuminemia < 3.1 mg/dL |  |  |  | 0.298 |
| No – n (%) | 55 (26.6%) | 20 (32.3%) | 35 (24.1%) |  |
| Yes – n (%) | 152 (73.4%) | 42 (67.7%) | 110 (75.9%) |  |
| Etiology |  |  |  | 0.526 |
| Atypical Bacteria – n (%) | 86 (32.8%) | 23 (29.5%) | 63 (34.2%) |  |
| Typical Bacteria – n (%) | 38 (14.5%) | 14 (17.9%) | 24 (13.0%) |  |
| Virus – n (%) | 138 (52.7%) | 41 (52.6%) | 97 (52.7%) |  |
