## Supplementary material for "A tool to distinguish viral from bacterial pneumonia": Table 2

**Table 2. Variables included in the score. Weight of the values should be added to obtain the total value of the score. Step 1 differentiates viral CAP from bacterial CAP (cut-off point, 11, see figure 5 for probabilities of bacterial CAP according punctuation). Step 2 differentiates, among those classified as bacterial CAP, typical bacterial CAP from atypical bacterial CAP (Cut-off point, 11.7, see figure 6 for probabilities of typical bacterial according punctuation). The result of the scores can be calculated quickly and easily in this online app:** <https://saradominguez-rodriguez.shinyapps.io/ValsDance_app/> (username: user; password: 0000).

**Accuracy of Step 1: Sensitivity, 97.3%. Specificity, 53.4%. The area under the receiver-operator curve (ROC) curve: 0.86.**

**Accuracy of Step 2: Sensitivity, 100%. Specificity, 60%. The area under the ROC curve, 0.97.**

| **Step 1 (viral <11 vs. bacterial >11)** | **Weight** |
| --- | --- |
| Age at admission >3 years | 10.6 |
| Zero Pneumococcal Conjugate Vaccine doses | 1.2 |
| Lack of work of breathing | 2.2 |
| Lack of wheezing | 1 |
| Temperature >37.7ºC | 1.3 |
| Consolidation on X-Ray | 5.5 |
| Hemoglobin >11 g/dL | 2.3 |
| Leukocytosis >15 000 cells/mm^3^ or leukopenia <4 000 cells/mm^3^ | 1.1 |
| Neutrophilia >10 000 cells/mm^3^ | 1.2 |
| C-reactive protein > 100 mg/L | 2.2 |
| **Step 2 (atypical bacteria <11.7 vs. atypical bacteria >11.7)** | **Weight** |
| Age at admission <3 years | 6.8 |
| Lack of cough | 3.0 |
| Lack of wheezing | 5.0 |
| Work of breathing | 5.8 |
| Hemoglobin <11 g/dL | 5.4 |
| Leukocytosis >15 000 cells/mm^3^ or leukopenia <4 000 cells/mm^3^ | 2.4 |
| Neutrophilia >10 000 cells/mm^3^ | 3.3 |
